# Perinatal Risk Factors Associated with Autism Spectrum Disorder

**DOI:** 10.64898/2026.03.12.26348254

**Authors:** Verónica L. Pantoja Silva, Vanessa P. Weinberger, Daniela Barriga, Nicole Garrido, Daniela Machuca, Natalia Salvadores

**Author notes:** Contributed equally (Verónica L. Pantoja Silva, Vanessa P. Weinberger).

## Abstract

Autism Spectrum Disorder (ASD) is a lifelong neurodevelopmental condition with multifactorial etiology, resulting from complex interactions between genetic susceptibility and environmental exposures. Although numerous studies have identified individual perinatal risk factors for ASD, most have examined these exposures in isolation, limiting understanding of how perinatal complications cluster and jointly influence neurodevelopment. Evidence from Latin America also remains scarce. This study aimed to identify multivariate perinatal risk patterns associated with ASD in a Chilean population, addressing gaps in regional representation and methodological approaches. We conducted a population-based analysis of mothers of children with and without ASD in Chile. A broad set of medical and psychosocial perinatal variables was jointly analyzed using multiple correspondence analysis (MCA) to characterize interrelated risk structures. MCA revealed a clear separation between ASD and non-ASD groups along the first dimension, suggesting that ASD diagnosis is embedded within structured perinatal patterns rather than isolated exposures. MCA-derived, stepwise, and LASSO-penalized logistic regression models were then compared. The most parsimonious model identified maternal vaginal bleeding during pregnancy, prenatal maternal stress or anxiety, and negative pregnancy intention or perception as the strongest factors jointly associated with increased odds of ASD, with a dose-response pattern observed for maternal stress. An unexpected inverse association with neonatal cyanosis may reflect enhanced medical surveillance and warrants cautious interpretation. These findings underscore the importance of integrated perinatal care addressing both obstetric and maternal mental health, and demonstrate the value of multivariate approaches for elucidating complex developmental risk pathways.

## Introduction

Autism Spectrum Disorder (ASD) is a lifelong neurodevelopmental condition characterized by persistent differences in social communication and interaction, alongside restricted, repetitive patterns of behavior and interests. Symptoms typically emerge in early childhood and can vary widely in severity and functional impact. ASD has gained increasing global attention due to its rising prevalence and its broad implications for educational systems, healthcare services, and social support structures. Recent meta-analytic evidence suggests that the global prevalence of ASD in children is approximately 0.77%, with considerable regional variability attributed to diagnostic practices and awareness^1^.

The etiology of ASD is multifactorial, involving complex interactions between genetic and environmental influences. Twin and family studies support a strong heritable component^2,3^, while environmental exposures during critical neurodevelopmental windows also contribute to ASD risk^4–6^. Among these, parental age^7–11^, prenatal medical conditions^4^, and maternal psychological stress^12–14^, have been consistently associated with increased ASD risk.

Perinatal complications such as fetal distress, umbilical cord problems, low Apgar scores, preterm birth, and low birth weight have all been reported in association with increased ASD risk across epidemiologic studies. A comprehensive meta-analysis examining over 60 perinatal and neonatal exposures identified several conditions, including fetal distress, birth trauma, and low birth weight, as being statistically linked to later ASD^15^, suggesting that compromised perinatal health may adversely influence neurodevelopmental outcomes. Subsequent meta-analyses and systematic reviews have additionally identified caesarean delivery, gestational age ≤36 weeks, breech or abnormal presentation, and hypertension-related perinatal conditions as associated with elevated ASD risk, although causal inference remains limited and heterogeneity across studies is substantial^16–18^. Retrospective cohort analyses further indicate that combined antepartum and intrapartum complications are associated with a 15-44% increased hazard of ASD compared to unexposed children, underscoring the potential cumulative impact of perinatal stressors^19^. However, findings across studies vary widely; while some perinatal factors achieve statistical significance in specific populations, others show inconsistent or modest effect. Recent work also highlights that perinatal risk may be influenced by broader familial and multigenerational characteristics, complicating causal interpretation^20,21^.

Despite substantial epidemiologic evidence linking individual perinatal factors to ASD risk, important gaps remain in the current literature. Most prior studies have evaluated perinatal exposures in isolation using univariable or contingency-table-based approaches, limiting the ability to capture the joint structure, clustering, and cumulative patterns of perinatal complications that may better reflect real-world clinical scenarios. Additionally, many analyses have relied on populations from North America or Europe, raising concerns about the generalizability of findings to underrepresented regions, including Latin America, where differences in healthcare systems, obstetric practices, sociodemographic factors, and population genetics may modify perinatal risk profiles. To date, no study has comprehensively examined perinatal risk factors for ASD using multivariate methods in a Chilean population. In the present study, we address these gaps by jointly analyzing a broad set of categorical perinatal variables using multiple correspondence analysis (MCA) combined with logistic regression, enabling the identification of global perinatal risk patterns and the estimation of associations with ASD while accounting for interrelated exposures. By providing the first population-based evaluation of perinatal risk profiles for ASD in Chile, this work contributes novel regional evidence and advances methodological approaches for understanding the complex perinatal determinants of autism.

## Methods

### Ethics approval

Study approval was obtained from the Scientific Ethics Committee of Universidad Mayor (No. 0530).

### Consent to participate

Informed consent was obtained from all individual participants included in the study.

### Participants

Participants were drawn from a private educational institution in Chile, operating virtually. The sample included 95 mothers of children diagnosed with ASD and 139 mothers of children without ASD. Mothers of children in early schooling stages were selected to minimize recall bias.

### Survey

The *Maternal Perinatal Risk Questionnaire*^22,23^ was used. This questionnaire is a structured self-report instrument that includes a set of risk factors that may hinder typical pregnancy development, as well as social factors that may influence the perinatal period. The questionnaire consists of 43 structured questions addressing various aspects related to the participants’ pregnancies. It included questions on maternal medical history, health conditions during pregnancy, complications during childbirth, and support received during the perinatal period, among other relevant factors (**Table 1**). We used questionnaire responses as data for comparing ASD children diagnosis.

**Table 1:** Variables and their corresponding levels analyzed in this study, after data curation.

| ID | Question/Variable | Level Details (number of cases in bold) |
| --- | --- | --- |
| Q01 | ASD diagnosis of the child | Q01_L1: diagnosed with ASD **(95)**<br>Q01_L2: not diagnosed with ASD **(139)** |
| Q02 | Sex of the child | Q02_L1: Female **(137)**<br>Q02_L2: Male **(97)** |
| Q1 | Weight of the mother before getting pregnant | Q1_L1: <40Kg **(19)**<br>Q1_L2: [40Kg-50Kg] **(35)**<br>Q1_L3: [51Kg-60Kg] **(59)**<br>Q1_L4: [61Kg-70Kg] **(93)**<br>Q1_L5: >71Kg **(28)** |
| Q2 | Height of the mother before getting pregnant | 1: <1,60m **(136)**<br>2: [1,60m-1,70m] **(77)**<br>3: >1,70m **(21)** |
| Q3 | Number of previous pregnancies before this child | 1: first pregnancy **(61)**<br>2: had previous children **(173)** |
| Q4 | Number of spontaneous abortions before this pregnancy | 1: no abortions **(169)**<br>2: previous abortions **(65)** |
| Q5 | Number of voluntary abortions before this pregnancy | 1: no abortions **(205)**<br>2: previous abortions **(29)** |
| Q6 | History of significant problems, delays, or disorders in any former child | 1: no conditions **(150)**<br>2: other children with conditions **(84)** |
| Q8 | Surgical interventions before/during this or previous pregnancies | 1: no intervention **(159)**<br>2: with intervention **(75)** |
| Q9 | Whether the mother used anticonception methods for avoiding this pregnancy | 1: no anticonception method **(133)**<br>2: hormonal methods **(48)**<br>3: barrier or DIU methods **(53)** |
| Q11 | Whether the mother suffered specified diseases before getting pregnant | 1: did not suffer any disease **(154)**<br>2: did suffer at least one of the diseases **(80)** |
| Q12 | Weight gained during pregnancy | 1: less than 4Kg **(28)**<br>2: [4Kg-7Kg] **(84)**<br>3: [8Kg-11Kg] **(55)**<br>4: [12Kg-15Kg] **(39)**<br>5: >15Kg **(28)** |
| Q14 | Vaginal bleeding during pregnancy | 1: Never **(70)**<br>2: Sometimes (not discerning whether at the beginning or at the end) **(136)**<br>3: frequently, during the entire pregnancy **(28)** |
| Q16 | Whether the mother smoked cigarettes during pregnancy and its frequency | 1: No **(197)**<br>2: Yes **(37)** |
| Q17 | Whether the mother consumed alcohol during pregnancy and its frequency | 1: No **(217)**<br>2: Yes **(17)** |
| Q19 | Frequency of nausea during pregnancy | 1: Never **(124)**<br>2: Sometimes **(80)**<br>3: All the time **(30)** |
| Q20 | Mother experienced edema during pregnancy | 1: Never **(76)**<br>2: Sometimes at the beginning **(90)**<br>3: Sometimes at the end **(24)**<br>4: All the time **(44)** |
| Q24 | Labor details (cesarean section) | 1: normal labor (not c-section) **(94)**<br>2: c-section **(140)** |
| Q25 | Fetal position during labor | 1: Correct position **(194)**<br>2: Incorrect position **(40)** |
| Q26 | Length of labor | 1: <3 hours **(189)**<br>2: [3-10 hours] **(24)**<br>3: >10 hours **(21)** |
| Q27 | Induced labor | 1: Not induced **(140)**<br>2: Induced **(94)** |
| Q31 | Infant weight at birth | 1: <1.5Kg **(10)**<br>2: [1.5Kg-2.5Kg] **(138)**<br>3: (2.5Kg-3.5Kg] **(77)**<br>4: >3.5Kg. **(9)** |
| Q32 | Length of pregnancy | 1: <28 weeks **(11)**<br>2: [28weeks-36weeks] **(48)**<br>3: [37weeks-41weeks] **(140)**<br>4: >41 weeks **(35)** |
| Q33 | Baby had bluish/purplish color at birth requiring medical care | 1: No **(111)**<br>2: Yes, in some parts **(24)**<br>3: Yes, all over the body **(99)** |
| Q34 | Special care for the born baby | 1: No **(142)**<br>2: Yes, she/he needed incubator **(45)**<br>3: Yes, she/he needed urgency care **(40)**<br>4: Yes, she/he needed surgery **(7)** |
| Q36 | Maternal stress/anxiety during pregnancy | 1: No, almost never **(106)**<br>2: Occasionally (1-2 per month) **(65)**<br>3: Frequently (1-2 per week) **(33)**<br>4: Very frequent (daily) **(30)** |
| Q37 | Maternal sadness, loneliness, or depression during pregnancy | 1: No, almost never **(107)**<br>2: Occasionally (1-2 per month) **(61)**<br>3: Frequently (1-2 per week) **(38)**<br>4: Very frequent (daily) **(28)** |
| Q39 | Overall quality of partner relationship during this pregnancy | 1: Normal relation **(187)**<br>2: Tense and/or complicated **(39)**<br>3: Single mother **(8)** |
| Q40 | Physical activity during pregnancy | 1: No, almost no physical work performed **(96)**<br>2: Some, but very mild **(54)**<br>3: Yes, only at the beginning of the pregnancy (until 3rd month) **(34)**<br>4: Yes, extending after the third month and almost until the end of pregnancy **(50)** |
| Q41 | Mental activity during pregnancy (exhaustive and/or with big responsibility) | 1: No, almost no exhaustive mental activity performed **(60)**<br>2: Yes, but very mild **(81)**<br>3: Yes, only at the beginning of the pregnancy (until 3rd month) **(44)**<br>4: Yes, only until the middle of the pregnancy (until 5th month) **(28)**<br>5: Yes, until very late or the end of the pregnancy **(21)** |
| Q42 | Overall experience of pregnancy | 1: relaxed and joyful **(80)**<br>2: happy, although i had some moments that i did not enjoy **(54)**<br>3: sometimes cheerful, sometimes depressed **(43)**<br>4: Very nervous, restless and distressed **(35)**<br>5: With great fear and distress **(22)** |
| Q43 | Intended pregnancy | 1: Yes, it was desired and planned **(105)** |
|  |  | 2: Yes, it was desired but not planned nor expected when it happened **(60)**<br>3: No, it was unpleasant news **(30)**<br>4: No, it was bad timing **(39)** |

### Data treatment

The questionnaire comprised 43 questions, which were coded as 13 ordinal and 30 nominal categorical variables. Each question (Q_i_,with i=1-43) considered different number of response levels (L_j_, with j ranging from 2 to 5 levels). Appendix A details the nomenclature and questionnaire coding used to construct this study database.

Data curation addressed count imbalances within levels. Specifically, we (i) merged levels when the number of counts or responses in one level was too low (<5 observations); and (ii) eliminated questions when count distributions were insufficient to define meaningful categories for analysis (e.g. all responses in one level) See Appendix A for details. Data curation details are addressed in **Appendix A**. After this curation, 31 of the original 43 perinatal variables were retained (**Table 1**).

### Analyses

We performed Multiple Correspondence Analysis (MCA) and logistic regressions to explore and test associations between perinatal conditions and ASD diagnosis in children. MCA was used as an exploratory multivariate technique to reduce model dimensionality and identify dominant patterns of association among variables and responses (i.e. ASD diagnosis). This approach decomposes the categorical dataset into orthogonal dimensions that sequentially maximize explained data inertia; as variables contribute unequally to those dimensions, MCA analysis highlights the joint response patterns that most strongly structure the data.

Different variable selection approaches were implemented to identify combinations of perinatal factors that are most strongly associated with ASD. Based on the MCA, we selected variables whose contributions or squared cosines (cos²) to the principal dimensions were at least twice (>2.5) the value expected under equal contribution. Hierarchical clustering of variables was then applied to this subset to reduce variable redundancy, retaining the variables with the highest squared loading. In addition to these MCA-informed models, we also applied stepwise (forward-backward) logistic regressions and regressions with LASSO penalization to identify best-fitting variable combinations. Model performance and parsimony were compared between models using explained percentage of data inertia and the Akaike Information Criterion (AIC). Models explaining a higher proportion of inertia and/or presenting lower AIC values were considered better representations of the data and used for the logistic regression model. To ensure convergence and minimize small-sample bias, we employed Firth’s penalized maximum-likelihood logistic regression.

All statistical analyses were performed in R software, using the packages *FactoMineR*, *factoextra*, *ClustOfVar*, *MASS*, *logistf* and *glmnet*. Data and R scripts are available at the Github repository “https://github.com/vanewi/ASD-analyses”.

## Results

The MCA (**Figure 1**) revealed that ASD diagnosis was primarily separated along the first principal component or first dimension (Dim1), with “individuals” (i.e. *mothers*) having children with ASD diagnosis significantly grouped to the right (Q01_L1 coordinates: (0.90/0.09), vtest for first dimension = 11.32, eta2 = 0.55), while mothers with children without ASD diagnosis grouped to the left (Q01_L2 coordinates: (-0.61/0.06, v.test for first dimension=-11.32, eta2=0.55). Indeed, cluster analyses using the k-means algorithm (with k=2 as hypothesis) confirmed that the mean of these two groups (children with ASD vs. no ASD diagnosis) are significantly different along the first dimension (t-test, coords Q01_L1∼ cluster, t = 27.70, df = 232, p-value < 2.2e-16. Mean group 1 (no ASD) -0.39, Mean group 2 (ASD) = 0.35). Moreover, cluster membership aligned significantly with diagnosis: 78.4% of individuals with no ASD fell in cluster 1 (109/139 individuals), while 98.9% of individuals with ASD fell in cluster 2 (94/95 individuals) (Pearson Chi2 with Yates correction, X-squared = 132.5, df = 1, p-value < 2.2e-16). All of the above indicates that mothers of children with and without ASD occupy significantly different regions of the MCA space, indicating underlying response patterns. We note, however, that Dim1 and Dim2 explain 8.4% and 6.5% of total inertia, respectively (14.9% cumulatively); while subsequent dimensions account for <4% of inertia (not shown).

**Figure 1:**
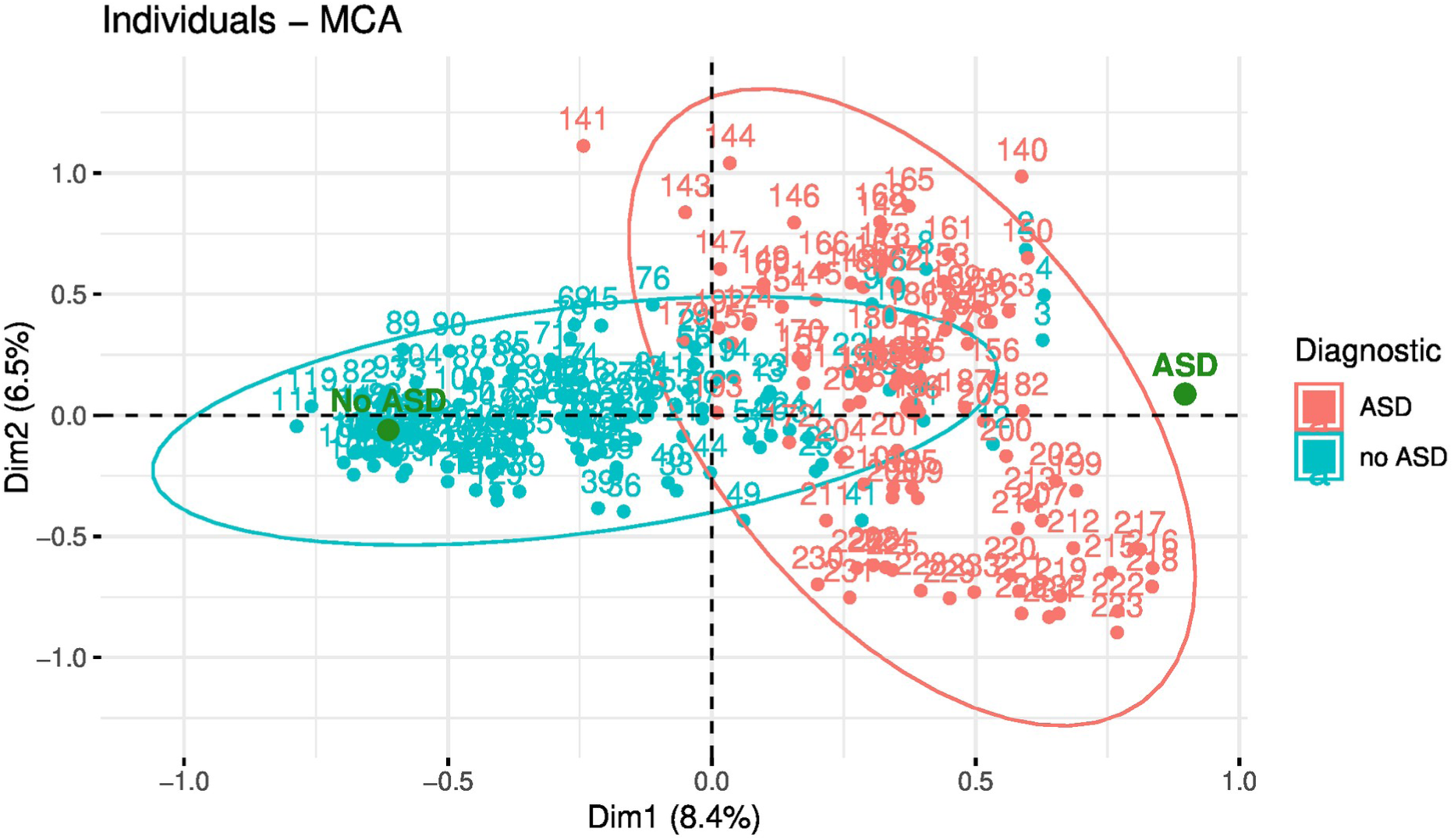
Multiple correspondence analysis (MCA) of individuals (i.e. mothers) based on the 31 curated perinatal variables. Points are colored by child diagnostic status (ASD vs. no ASD); green markers indicate the projected positions of the diagnostic categories.

Based on the ASD diagnostic pattern observed along the first MCA dimension, we investigated the variables that most associated to both and only the first principal component dimension, according to their squared cosine (cos^2^) and contributions. To minimize redundancy among highly correlated variables, we also performed hierarchical clustering on those selected variables. Four logistic regression models were suggested by these methods (models i-iv). Models v (lasso simple) and vi (lasso severe) were derived from variables selected through simple and strong LASSO-penalized logistic regressions, respectively. Stepwise forward–backward regression analyses did not identify alternative variable combinations providing a better fit to the data. All models are detailed in **Table 2**.

**Table 2:** Models tested for ASD diagnostic in children. These models are created from a subset of the original 31 perinatal variables. Details of variable selection are indicated in methods and along the text. Details about the percentage of data inertia explained in the first principal component of an MCA (%Dim1), and the second principal component of an MCA (%Dim2) are detailed, and also their accumulated percentage. The AIC is also shown, based on a logistic regression.

| Model | Variables Selected in the Model | %Dim1 | %Dim2 | %Accumulated | AIC |
| --- | --- | --- | --- | --- | --- |
| **(o) all** | Q02, Q1, Q2, Q3, Q4, Q5, Q6, Q8, Q9, Q11, Q12, Q14, Q16, Q17, Q19, Q20, Q24, Q25, Q26, Q27, Q31, Q32, Q33, Q34, Q36, Q37, Q39, Q40, Q41, Q42, Q43 | 8.4 | 6,5 | 14.9 | 321.02 |
| **(i) dim1dim2** | Q1, Q3, Q4, Q9, Q11, Q14, Q25, Q27, Q33, Q34, Q36, Q37, Q40, Q41, Q42, Q43 | 11.17 | 8.16 | 19.33 | 321.02 |
| **(ii) dim1** | Q1, Q9, Q11, Q14, Q33, Q34, Q36, Q37, Q42, Q43 | 14.02 | 6.25 | 20.27 | 318.90 |
| **(iii) dim1dim2 + cluster** | Q1, Q3, Q25, Q27, Q33, Q34, Q41 | 14.44 | 12.10 | 26.54 | 316.88 |
| **(iv) dim1+cluster** | Q9, Q14, Q33, Q34, Q36, Q43 | 17.89 | 10.58 | 28.47 | 313.33 |
| **(v) lasso simple** | Q1, Q3, Q4, Q11, Q14, Q25, Q33, Q34, Q36, Q37, Q40, Q41, Q42, Q43 | 11.38 | 8.31 | 19.68 | 321.00 |
| **(vi) lasso severe** | Q1, Q3, Q4, Q11, Q14, Q25, Q33, Q36, Q37, Q40, Q41, Q42, Q43 | 11.87 | 8.54 | 20.41 | 318.77 |

Model i (dim1dim2) included the variables that most strongly associated with both MCA dimensions. This model comprised 16 of the 31 perinatal variables (Q1, Q3, Q4, Q9, Q11, Q14, Q25, Q27, Q33, Q34, Q36, Q37, Q40, Q41, Q42, Q43), and increased the cumulative inertia of the first two dimensions to 19.33%, with Dim1 explaining 11.17%. Model ii (dim1) retained only the variables most strongly associated with the first dimension, resulting in 10 variables (Q1, Q9, Q11, Q14, Q33, Q34, Q36, Q37, Q42, Q43) and explaining 20.27% of the total inertia in the first two components, of which 14.02% corresponded to Dim1. Model iii (dim1dim2 + cluster) retained 7 perinatal variables (Q1, Q3, Q25, Q27, Q33, Q34, Q41), increasing the inertia explained by the first two dimensions to 26.54% (Dim1 = 14.44%). Model iv (dim1 + cluster) included 6 perinatal variables (Q9, Q14, Q33, Q34, Q36, Q43) and accounted for 28.47% of total inertia (Dim1 = 17.89%).

Finally, Models v (lasso simple) and vi (lasso severe) increased the percentage of inertia explained compared to the original MCA with all 31 perinatal variables. Model v included 14 of the 16 variables from Model i (Q1, Q3, Q4, Q11, Q14, Q25, Q33, Q34, Q36, Q37, Q40, Q41, Q42, Q43), explaining 19.68% of total inertia, with 11.38% explained by Dim1. Model vi comprised 13 variables (Q1, Q3, Q4, Q11, Q14, Q25, Q33, Q36, Q37, Q40, Q41, Q42, Q43), explaining 20.41% of total inertia and 11.87% by Dim1.

Across all evaluated models (i-vi), the ASD diagnostic pattern remained consistent along the first dimension, showing a consistent and statistically significant separation between the two groups as observed in the full-variable MCA (t-tests comparing cluster centroids and Pearson χ² tests with Yates correction; all p < 1e-07, details not shown).

Based on the percentage of inertia explained by the first two MCA dimensions and AIC values, model iv (dim1 + cluster) was identified as the best-fitting model. This model (i) explained one of the highest proportions of data inertia (28.47%) and (ii) exhibited the lowest AIC value (315.71) among all models (**Table 2**). We therefore evaluated the effect of the variables included in this model on the probability of diagnosing children with ASD using logistic regression (**Table 3**).

**Table 3.**
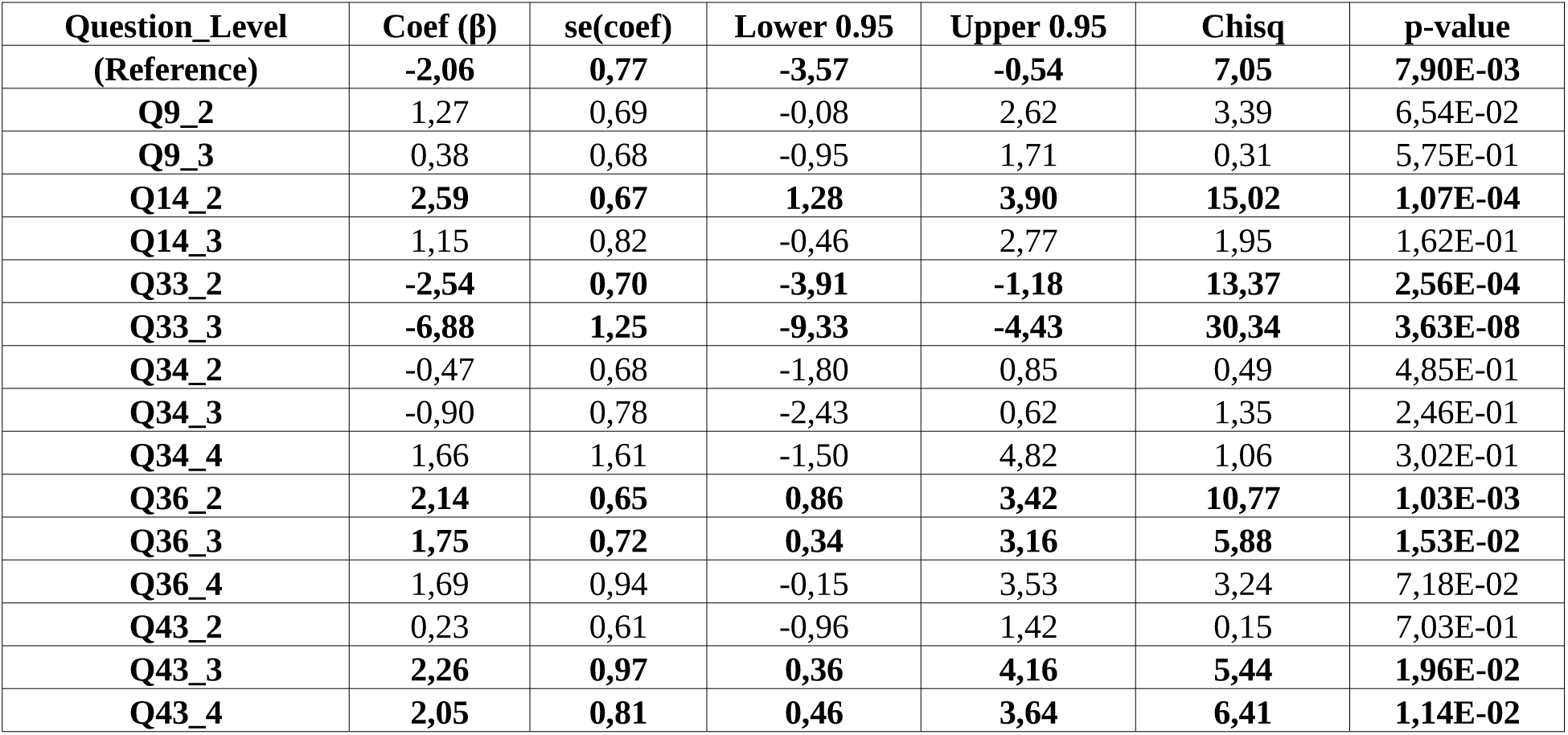
Logistic regression results for model iv (dim1 + cluster), showing predictors associated with the likelihood of having a child with ASD. Odds ratios (OR = exp(Coef)) indicate the direction and magnitude of each effect; significant predictors (p < 0.05) are in bold.

We start identifying that model iv was globally significant (Likelihood Ratio test = 217.85, df = 15, p < 1e-07; Wald test = 57.29, df = 15, p < 1e-07), indicating that the variables jointly improved the fit compared to the null model. In this model, reference women are mothers who did not use any anticonception method for avoiding the pregnancy (Q9_L1), never experienced vaginal bleeding during the pregnancy (Q14_L1), their child did not show a bluish/purplish color at birth that required medical care (Q33_L1) and her born baby did not require any special care (Q34_L1), she did not experience stress or anxiety during pregnancy (Q36_L1) and her pregnancy was intended and desired (Q43_L1).

The logistic regression model showed that the reference or intercept is significant (β₀ = -2.06, CI 95%= [-3.57/-0.54], p-value = 0.0079), indicating that under baseline conditions, the probability of having a child with ASD (11.3%) differs significantly from a random expectation of 50% (**Table 3**). However, under this model, the probability of having a child with ASD decreased when:

(i) The baby presented partial bluish/purplish coloration at birth, requiring medical care (Q33_2). In this case, the odds decreased by 0.08 relative to the reference, and the probability dropped to 1% (β_33_2_ = -2.54, CI 95% [-3.91, -1.18], p-value = 2.56x10^-4^). Moreover, if the baby exhibited complete bluish/purplish coloration at birth, requiring medical care (Q33_3), the odds reduced even more, by 0.001, and the probability dropped to nearly 0% (0,01%) (β_33_3_ = -6.88, CI 95% [-9.33, -4.43], p-value = 3.63x10^-8^).

Conversely, the probability of having a child with ASD diagnosis increased when:

(i) The mother experienced vaginal bleeding at some stage of pregnancy (Q14_2), with odds increasing 13.3-fold and probability rising to 63.0% (β₁₄₂ = 2.59, 95% CI [1.28, 3.90], p = 1.07×10⁻⁴).
(ii) The mother experienced occasional stress or anxiety during pregnancy (Q36_2), increasing odds 8.5-fold and probability to 52.1% (β₃₆₂ = 2.14, 95% CI [0.86, 3.42], p = 1.03×10⁻³). When stress was frequent (Q36_3), odds increased 5.75-fold, and the probability reached 42.4% (β₃₆₃ = 1.75, 95% CI [0.34, 3.16], p = 1.53×10⁻²).
(iii) The pregnancy was perceived as unpleasant news (Q43_3), increasing odds 9.6-fold and probability to 55.1% (β₄₃₃ = 2.26, 95% CI [0.36, 4.16], p = 1.96×10⁻²). When the pregnancy was considered badly timed (Q43_4), odds increased 7.8-fold and probability reached 49.9% (β₄₃₄ = 2.05, 95% CI [0.46, 3.64], p = 1.14×10⁻²).

All other variables did not significantly affect the odds or probabilities of having a child diagnosed with ASD.

## Discussion

In this study, we jointly analyzed a broad set of perinatal variables using multivariate methods to characterize perinatal risk profiles associated with ASD in a Chilean population. Our findings demonstrate a clear separation between mothers of children with and without ASD along the first dimension of a multiple correspondence analysis (MCA), indicating that ASD diagnosis is embedded within a structured group of perinatal experiences rather than isolated exposures. This supports the growing view that ASD risk is influenced by cumulative and interacting perinatal factors.

Among the six evaluated models, the most parsimonious MCA-informed model (model iv) provided the best fit to the data and identified a limited set of perinatal variables (6 main variables) that jointly explained a substantial proportion of data inertia (28.47%) and significantly predicted ASD diagnosis. Notably, maternal vaginal bleeding during pregnancy, maternal stress or anxiety, and negative pregnancy intention or perception emerged as the strongest risk-associated factors. These findings are consistent with previous epidemiological studies linking prenatal stress, obstetric complications, and adverse psychosocial contexts to elevated ASD risk^24–26^. Maternal vaginal bleeding has been previously associated with increased risk of neurodevelopmental disorders, including ASD, potentially reflecting placental dysfunction, inflammation, or hypoxic processes during critical periods of fetal brain development^27–29^. The strong association observed in our logistic regression analysis aligns with this literature and suggests that even transient obstetric complications may signal broader gestational instability relevant to neurodevelopmental outcomes. Maternal stress and anxiety during pregnancy also showed a robust association with ASD diagnosis. These results support prior evidence that prenatal maternal psychological distress may influence fetal neurodevelopment through hypothalamic-pituitary-adrenal axis dysregulation, altered cortisol exposure, and immune-mediated mechanisms^30–33^. The dose-related pattern observed in our study further strengthens the plausibility of this association. Interestingly, negative pregnancy intention or perception, specifically pregnancies perceived as unpleasant news or poorly timed, was associated with significantly increased ASD odds. While pregnancy intention has been less frequently examined in ASD research, similar associations have been reported in studies linking unintended pregnancy to adverse birth outcomes and neurodevelopmental vulnerability^34–40^. These findings suggest that psychosocial context and maternal emotional response to pregnancy may exert long-term developmental influences, potentially mediated by stress pathways or reduced prenatal care engagement. Conversely, the finding that neonatal cyanosis requiring medical care was associated with reduced ASD odds was unexpected and contrasts with much of the existing literature linking perinatal hypoxia to neurodevelopmental risk^41–45^. One possible explanation is that this variable may reflect heightened medical surveillance rather than hypoxia severity per se.

The statistically significant intercept observed in Model iv indicates that, even under the reference conditions characterized by the absence of the perinatal factors evaluated and by generally favorable pregnancy and birth histories, there remains a meaningful baseline probability of ASD diagnosis. In practical terms, this suggests that children of mothers who did not present detectable perinatal risk according to the variables examined may still have an elevated likelihood of ASD, pointing to the influence of factors beyond the perinatal domain assessed in this study. These influences may involve broader environmental, psychosocial, familial, or postnatal contexts that were not captured by the questionnaire and are increasingly recognized as contributing to the complex and multifactorial nature of ASD^18,46^. Within this broader perspective, the finding that neonatal bluish or purplish coloration requiring medical attention was associated with a substantial reduction in ASD odds becomes particularly striking, as it challenges the assumption that perinatal complications invariably increase neurodevelopmental risk. Rather than suggesting a direct protective biological effect, this pattern may reflect contextual factors such as closer medical monitoring, earlier clinical attention, or different care trajectories, highlighting the importance of interpreting perinatal indicators within an environmental and developmental framework that extends beyond isolated biomedical events.

Methodologically, this study advances ASD perinatal research by integrating multiple correspondence analysis with logistic regression, enabling identification of multivariate risk patterns while mitigating collinearity and small-sample bias. This approach addresses limitations of prior univariate analyses and may better reflect real-world clustering of perinatal exposures. Importantly, this work provides novel evidence from Chile, a region underrepresented in ASD epidemiology, contributing to improved global generalization. Several limitations should be acknowledged. The retrospective, self-report design may be subject to recall bias, particularly for emotionally salient events. The sample was drawn from a single educational institution, potentially limiting external validity. Additionally, causal inference cannot be established due to the observational nature of the study, and stronger analyses could not be performed due to the categorical and unbalanced nature of the questionnaire. We recommend revisiting its questions for addressing other robust statistical analyses.

## Conclusion

Our findings indicate that ASD risk in this Chilean cohort is associated with specific patterns of perinatal medical and psychosocial factors, particularly maternal bleeding, stress, and pregnancy perception. These results underscore the importance of integrated perinatal care that includes both obstetric monitoring and maternal mental health support and highlight the value of multivariate analytical frameworks for elucidating complex developmental risk pathways.

## Data Availability

All data produced are available online at https://github.com/vanewi/ASD-analyses

## Statements and Declarations

### Competing Interests

The authors have no relevant financial or non-financial interests to disclose.

### Author contributions

V.P. D.B., N.G., and D.M. conceptualized the study, designed the methodology, and collected the data. V.P.W. performed the data analyses and drafted the manuscript. N.S. supervised the study and contributed to manuscript writing. All authors reviewed and approved the final manuscript.

### Funding sources

This research did not receive any specific grant from funding agencies in the public, commercial, or not-for-profit sectors.

## APPENDIX A

### Data Treatment

Details of the modifications made from original survey for the statistical analyses performed in this study.

| ID | Question/Variable | Type | Changes made for this study | Number of Levels | Level Details (number of cases in bold) |
| --- | --- | --- | --- | --- | --- |
| Q01 | ASD diagnosis of the child | Nominal | None | 2 | Q01_L1: diagnosed with ASD **(95)**<br>Q01_L2: not diagnosed with ASD **(139)** |
| Q02 | Sex of the child | Nominal | None | 2 | Q02_L1: Female **(137)**<br>Q02_L2: Male **(97)** |
| Q1 | Weight of the mother before getting pregnant | Ordinal | We merged the last two levels of the original survey to address sparse data in one level (<5 cases) | 5 | Q1_L1: <40Kg **(19)**<br>Q1_L2: [40Kg-50Kg] **(35)**<br>Q1_L3: [51Kg-60Kg] **(59)**<br>Q1_L4: [61Kg-70Kg] **(93)**<br>Q1_L5: >71Kg **(28)** |
| Q2 | Height of the mother before getting pregnant | Ordinal | We merged the first two and the last two levels of the original survey to address sparse data in one level (<5 cases) | 3 | 1: <1,60m **(136)**<br>2: [1,60m-1,70m] **(77)**<br>3: >1,70m **(21)** |
| Q3 | Number of previous pregnancies before this child | Ordinal | We merged the original levels of the survey and created a binary variable | 2 | 1: first pregnancy **(61)**<br>2: had previous children **(173)** |
| Q4 | Number of spontaneous abortions before this pregnancy | Ordinal | We merged the original levels of the survey and created a binary variable | 2 | 1: no abortions **(169)**<br>2: previous abortions **(65)** |
| Q5 | Number of voluntary abortions before this pregnancy | Ordinal | We merged the original levels of the survey and created a binary variable | 2 | 1: no abortions **(205)**<br>2: previous abortions **(29)** |
| Q6 | History of significant problems, delays, or disorders in any former child | Nominal | We did not analyze the type of problem described, only whether the were former children with conditions | 2 | 1: no conditions **(150)**<br>2: other children with conditions **(84)** |
| Q7 | Situations experimented in previous pregnancy (related to pregnancy problems) | Nominal | We excluded this question because the data were insufficient to define meaningful categories for analysis. | 0 | Not Applicable NA |
| Q8 | Surgical interventions before/during this or previous pregnancies | Nominal | We merged the original levels of the survey and created a binary variable | 2 | 1: no intervention **(159)**<br>2: with intervention **(75)** |
| Q9 | Whether the mother used anticonception methods for avoiding this pregnancy | Nominal | We excluded the last level of original survey due to lack of data, and merged original levels 3 and 4 of the survey to address sparse data | 3 | 1: no anticonception method **(133)**<br>2: hormonal methods **(48)**<br>3: barrier or DIU methods **(53)** |
| Q10 | Age when getting pregnant | Ordinal | We excluded this question because all mothers were in the same level (24-27 years) | 0 | Not Applicable NA |
| Q11 | Whether the mother suffered specified diseases before getting pregnant | Nominal | We merged the original levels of the survey and created a binary variable | 2 | 1: did not suffer any disease **(154)**<br>2: did suffer at least one of the diseases **(80)** |
| Q12 | Weight gained during pregnancy | Ordinal | We merged the last two levels of the original survey to address sparse data in one level (<5 cases) | 5 | 1: less than 4Kg **(28)**<br>2: [4Kg-7Kg] **(84)**<br>3: [8Kg-11Kg] **(55)**<br>4: [12Kg-15Kg] **(39)**<br>5: >15Kg **(28)** |
| Q13 | Month of pregnancy at first medical visit | Ordinal | We excluded this question because most mothers were in the same level (first month) | 0 | Not Applicable NA |
| Q14 | Vaginal bleeding during pregnancy | Nominal | We merged original levels 2 and 3 to address sparse data in one level (<5 cases) | 3 | 1: Never **(70)**<br>2: Sometimes (not discerning whether at the beginning or at the end) **(136)**<br>3: frequently, during the entire pregnancy **(28)** |
| Q15 | Remedies taken during pregnancy | Nominal | We excluded this question because the data were insufficient to define meaningful categories for analysis (most women did not take remedies) | 0 | Not applicable NA |
| Q16 | Whether the mother smoked cigarettes during pregnancy and its frequency | Nominal | We merged the original levels of the survey and created a binary variable due to the distribution of the data | 2 | 1: No **(197)**<br>2: Yes **(37)** |
| Q17 | Whether the mother drank alcohol during pregnancy and its frequency | Nominal | We merged the original levels of the survey and created a binary variable due to the distribution of the data | 2 | 1: No **(217)**<br>2: Yes **(17)** |
| Q18 | Whether the mother used illegal drugs during the pregnancy and its frequency | Nominal | We excluded this question because the data were insufficient to define meaningful categories for analysis (most women did not take illegal drugs) | 0 | Not applicable NA |
| Q19 | Frequency of nausea during pregnancy | Nominal | We merged level 2 and 3 of original survey due to sparse data | 3 | 1: Never **(124)**<br>2: Sometimes **(80)**<br>3: All the time **(30)** |
| Q20 | Mother experienced edema during pregnancy | Nominal | None | 4 | 1: Never **(76)**<br>2: Sometimes at the beginning **(90)**<br>3: Sometimes at the end **(24)**<br>4: All the time **(44)** |
| Q21 | Mother experienced any physical accidents or injuries | Nominal | We excluded this question because no | 0 | Not applicable NA |
|  | during pregnancy |  | mother experienced any accident or injury |  |  |
| Q22 | Diseases experienced during the pregnancy | Nominal | We excluded this question because the data were insufficient to define meaningful categories for analysis (not many woman experienced the diseases tested) | 0 | Not applicable NA |
| Q23 | Anesthesia taken during labor | Nominal | We excluded this question because the data were insufficient to define meaningful categories for analysis (most women used epidural) | 0 | Not applicable NA |
| Q24 | Labor details (cesarean section) | Nominal | We merged the original levels of the survey and created a binary variable due to the distribution of the data | 2 | 1: normal labor (not c-section) **(94)**<br>2: c-section **(140)** |
| Q25 | Fetal position during labor | Nominal | We merged the original levels of the survey and created a binary variable due to the distribution of the data | 2 | 1: Correct position **(194)**<br>2: Incorrect position **(40)** |
| Q26 | Length of labor | Ordinal | We merged levels 1 and 2; levels 3 and 4, and leveles 5-7 of original survey to address sparse data | 3 | 1: <3 hours **(189)**<br>2: [3-10 hours] **(24)**<br>3:>10 hours **(21)** |
| Q27 | Induced labor | Nominal | We merged the original levels of the survey and created a binary variable due to the distribution of the data. | 2 | 1: Not induced **(140)**<br>2: Induced **(94)** |
| Q28 | Forceps used in labor | Nominal | We excluded this question because the data were insufficient to define meaningful categories for analysis (most women did not used forceps) | 0 | Not applicable NA |
| Q29 | Difficulties during labor | Nominal | We excluded this question because the data were insufficient to define meaningful categories for analysis (most women did not show complications) | 0 | Not applicable NA |
| Q30 | Multiple birth | Ordinal | We excluded this question because the data were insufficient to define meaningful categories for analysis<br>(most women did not have multiple birth) | 0 | Not applicable NA |
| Q31 | Infant weight at birth | Ordinal | We merged the last two levels of the original survey to address sparse data in one level (<5 cases) | 4 | 1: <1.5Kg **(10)**<br>2: [1.5Kg-2.5Kg] **(138)**<br>3:(2.5Kg-3.5Kg) **(77)**<br>4: >3.5Kg. **(9)** |
| Q32 | Lenght of pregnancy | Ordinal | None | 4 | 1: <28 weeks **(11)**<br>2: [28weeks-36weeks] **(48)**<br>3: [37weeks-41weeks] **(140)**<br>4: >41 weeks **(35)** |
| Q33 | Baby had bluish/purplish color at birth requiring medical care | Nominal | We merged the answer “not sure” as “no”. | 3 | 1: No **(111)**<br>2: Yes, in some parts **(24)**<br>3: Yes, all over the body **(99)** |
| Q34 | Special care for the born baby | Nominal | We merged the answer “not sure” as “no”. | 4 | 1: No **(142)**<br>2: Yes, she/he needed incubator **(45)**<br>3: Yes, she/he needed urgency care **(40)**<br>4: Yes, she/he needed surgery **(7)** |
| Q35 | Apgar score | Ordinal | We excluded this question because the data were insufficient to define meaningful categories for analysis (most women did not remember the score) | 0 | Not applicable NA |
| Q36 | Maternal stress/anxiety during pregnancy | Nominal | None | 4 | 1: No, almost never **(106)**<br>2: Occasionally (1-2 per month) **(65)**<br>3: Frequently (1-2 per week) **(33)**<br>4: Very frequent (daily) **(30)** |
| Q37 | Maternal sadness, loneliness, or depression during pregnancy | Nominal | None | 4 | 1: No, almost never **(107)**<br>2: Occasionally (1-2 per month) **(61)**<br>3: Frequently (1-2 per week) **(38)**<br>4: Very frequent (daily) **(28)** |
| Q38 | Reaction to knowing baby’s sex before birth | Nominal | We excluded this question because the data were insufficient to define meaningful categories for analysis | 0 | Not applicable NA |
| Q39 | Overall quality of partner relationship during this pregnancy | Nominal | We merged level 2 and 3 of original survey to address sparse data in one level (<5 cases) | 3 | 1: Normal relation **(187)**<br>2: Tense and/or complicated **(39)**<br>3: Single mother **(8)** |
| Q40 | Physical activity during pregnancy | Nominal | We excluded level 6 of original survey for not having data, and merged level 4 and 5 to address | 4 | 1: No, almost no physical work performed **(96)**<br>2: Some, but very mild **(54)**<br>3: Yes, only at the beginning of |
|  |  |  | sparse data in one level (<5 cases) |  | the pregnancy (until 3rd month) **(34)**<br>4: Yes, extending after the third month and almost until the end of pregnancy **(50)** |
| Q41 | Mental activity during pregnancy (exhaustive and/or with big responsibility) | Nominal | We merged level 5 and 6 of original survey to address sparse data in one level (<5 cases) | 5 | 1: No, almost no exhaustive mental activity performed **(60)**<br>2: Yes, but very mild **(81)**<br>3: Yes, only at the beginning of the pregnancy (until 3rd month) **(44)**<br>4: Yes, only until the middle of the pregnancy (until 5th month) **(28)**<br>5: Yes, until very late or the end of the pregnancy **(21)** |
| Q42 | Overall experience of pregnancy | Nominal | We excluded level 6 because of no data | 5 | 1: relaxed and joyful **(80)**<br>2: happy, although i had some moments that i did not enjoy **(54)**<br>3: sometimes cheerful, sometimes depressed **(43)**<br>4: Very nervous, restless and distressed **(35)**<br>5: With great fear and distress **(22)** |
| Q43 | Inteded pregnancy | Nominal | None | 4 | 1: Yes, it was desired and planned **(105)**<br>2: Yes, it was desired but not planned nor expected when it happened **(60)**<br>3: No, it was an unpleasant news **(30)**<br>4: No, it was a bad timing **(39)** |

## Notes

### Competing Interest Statement

The authors have declared no competing interest.

### Author Declarations

Scientific Ethics Committee of Universidad Mayor (No. 0530).

